# Efficacy and safety of newer antibiotics versus generic antibiotics for hospital-acquired bacterial pneumonia and ventilator-associated bacterial pneumonia: a systematic review and meta-analysis of randomized controlled trials

**DOI:** 10.64898/2026.02.11.26345978

**Authors:** Anh T. K. Nguyen, David L. Paterson, Yin Mo, Yukiko Ezure, Pham Thanh Duy, C. Louise Thwaites

## Abstract

**Background:** Hospital-acquired bacterial pneumonia (HABP) and ventilator-associated bacterial pneumonia (VABP), particularly those caused by multi-drug resistant organisms (MDROs), often require newer antibiotic treatment. The efficacy and safety of newer antibiotics compared to generic antibiotics in randomized controlled trials (RCTs) have not been evaluated before.

**Methods:** In this systematic review, we searched RCTs in the United States National Library of Medicine (PubMed), Cochrane Central Register of Controlled Trials (CENTRAL), Scopus, Ovid MEDLINE, Clinical Trials.gov and Google Scholar databases published between 2013 and 2025. The primary efficacy endpoint was 28-day all-cause mortality. Secondary efficacy endpoints were clinical and microbiological response. Safety endpoint was nephrotoxicity.

**Results:** We identified eight eligible RCTs involving 2,881 patients (1,450 patients treated with newer antibiotics and 1,431 patients treated with generic antibiotics) with HABP/VABP. The meta-analysis did not reveal any significant differences between newer and generic antibiotics for all-cause mortality at day 28 (risk ratio (RR) 0.97, 95% confidence interval (CI) 0.72-1.30), clinical response (RR 1.04, 95%CI 0.93-1.17), and microbiological response (RR 1.05, 95%CI 0.89-1.24). However, newer antibiotics showed significant lower occurrences of nephrotoxicity compared to colistin component (RR 0.30, 95%CI 0.11-0.79). In subgroup analysis, newer antibiotic regimens demonstrated significant improvement in microbiological eradication of carbapenem-resistant Gram-negative bacilli (RR 1.50, 95%CI 1.18-1.90).

**Conclusions:** Newer antibiotics showed similar efficacy and safety in treating HABP/VABP compared to generic drugs. The superiority in microbiological eradication of carbapenem-resistant Gram-negative bacilli of newer antibiotics could suggest that future trials should be targeted for those patients to improve understanding of their therapeutic use and pathophysiology of these conditions.

**Key points:** Newer antibiotics, despite broader antimicrobial coverage, have not significantly outperformed generic comparators in terms of 28-day all-cause mortality, clinical, or microbiological response in patients with Gram-negative HABP/VABP. This may reflect limitations in current trial designs focused primarily on regulatory approval.

## INTRODUCTION

In the intensive care unit (ICU), hospital-acquired bacterial pneumonia (HABP), including ventilator-associated bacterial pneumonia (VABP) are the most common nosocomial infections. The point prevalence of HABP is 21.8% among 504 healthcare-associated infections [1]. In the USA, VABP represents the commonest nosocomial infection in critically ill patients, with incidence ranging from 6.1 to 28.5 episodes per 1,000 ventilator-days [2]. Importantly, the mortality associated with VABP ranges from 30% to 50% from surveillance and cohort studies, making it the most burdensome hospital-acquired infection in the ICU [2,3].

Both HABP and VABP are caused by a wide spectrum of bacterial pathogens. Infections due to aerobic Gram-negative bacilli are commonest, particularly *Klebsiella pneumoniae, Acinetobacter baumannii*, and *Pseudomonas aeruginosa* [4,5]. Rising Gram-negative pathogens resistance to antibiotics is widely reported as a major threat to ICU care globally [6–9]. The presence of MDR pathogens typically leads to delay in effective antibiotic therapy administration which is associated with increasing mortality [6,10–12]. A systemic analysis of antimicrobial resistant bacteria in 2019 reported the highest burden of disease in lower respiratory infections caused by pathogen-drug resistant combination, with the total of 1.5 million deaths. The three priority bacterial pathogens each caused 38,000-58,000 deaths: carbapenem-resistant *P aeruginosa*, carbapenem-resistant *K pneumoniae, and* carbapenem-resistant *A. baumannii* [13,14].

Due to the increasing prevalence of extended-spectrum β-lactamase-producing, and especially carbapenemases among the predominant HABP/VABP pathogens, only a few generic treatment options are available, particularly in low- and middle-income countries (LMICs). Over the last decade, five newer antibiotics have been produced and approved by the USA’s Food and Drug Administration to treat HABP/VABP, including ceftazidime-avibactam, cefiderocol, imipenem-cilastatin-relebactam, sulbactam-durlobactam and ceftolozane-tazobactam. These newer antibiotic regimens are not easily accessible in LMICs but potentially are a key component to improve treatment and outcomes for patients with HABP/VABP [15]. Nevertheless, the added cost burden associated including them as therapeutic options in these countries requires robust justification.

Despite this promise, the efficacy and safety of newer antibiotic therapies in the treatment of HABP and VABP has not been systematically summarized and analyzed from randomized controlled trials. Therefore, the aim of this systematic review and meta-analysis was to assess the efficacy and safety of newer antibiotics compared to generic antibiotic therapy for patients with HABP/VABP.

## METHODS

This paper adheres to the Preferred Reporting Items for Systematic reviews and Meta-Analyses (PRISMA) statement for reporting systematic reviews and meta-analysis 2020 and Assessing the methodological quality of systematic reviews (AMSTAR) 2 guideline [16,17] (see Supplementary Tables 1 and 2). The protocol of this study was registered with the International Prospective Register of Systematic Reviews (PROSPERO) on 13 November 2023 (registration number CRD42023476481).

### Search Strategy

On 7 September 2023, we conducted a systematic literature search in electronic databases, including PubMed, Cochrane Central Register of Controlled Trials (CENTRAL), Scopus and Ovid MEDLINE. We manually screened the reference list of included trials to identify additional articles. Additionally, we searched Clinical Trials (Home | ClinicalTrials.gov) and Google Scholar to ascertain completed and ongoing trials. Our search was limited to data published between 2013 and 2023. We updated the search strategy in January 2025 as there was a new trial published in December 2024. The literature was restricted to English only. The search strategies were discussed among the research team and the details are listed in Supplementary Text 1.

### Study Selection

Two authors (A.N. and D.P.) performed independent screening of the titles, abstracts and full text of selected articles to determine their eligibility according to the inclusion and exclusion criteria (see Supplementary Text 2). Generally, eligible trials were all available randomized controlled trials that compared newer antibiotics with generic antibiotics for the treatment of adult with HABP/VABP. Therefore, trials including placebo controls, study populations with community-acquired pneumonia and healthy volunteers were excluded.

### Assessment of the studies’ quality and risk of bias

RCTs were evaluated according to the Consolidated Standards of Reporting Trials (CONSORT) Statement of quality assessment [18]. The *Cochrane Collaboration’s tool for assessing risk of bias in randomised trials* (RoB 2, 2019) was used to revise the quality of included trials [19]. We evaluated the risk of bias in five distinct domains (A. randomization process, B. deviations from intended interventions, C. missing outcome data, D. measurement of outcome and E. selecting the reported results). The risk-of-bias judgments for each domain are “low risk”, “some concerns” or “high risk” according to the response of one or more signalling questions.) Then, an overall judgment for risk of bias in individual trials was resulted. Full details are shown in Supplementary Tables 3 and 4.

### Data Extraction and Outcome Measures

Relevant data from eligible studies were extracted by following the *Cochrane Handbook for Systematic Reviews of Interventions* version 6.4 (updated August 2023) [20], including: (i) Name of RCT, first author and publication year; (ii) study population including sample size and type of intent-to-treat population; (iii) characteristics of the participants, including mean age, sex, ventilation, ICU admission, renal function and Acute Physiology and Chronic Health Evaluation (APACHE) II score; (iv) newer and generic antibiotics, including component, number of participants, duration of antibiotics and (v) baseline low respiratory tract (LRT) pathogens. Any conflict was resolved by discussion between all reviewers and/or seeking advice from co-authors. To ensure the reliability of data, the data were extracted by two reviewers, independently.

The primary efficacy outcome was day 28 all-cause mortality. The safety outcome was measured by the proportion of patients suffered from nephrotoxicity which was defined as the increase of serum creatinine at different values according to the renal function of patients at baseline. Secondary efficacy outcomes included assessments of clinical and microbiological responses. Clinical response was defined as clinical cure or favorable clinical response at the visits. Definition of microbiological response comprised microbiological eradication and favorable microbiological response during the study period (details in Supplement Table 5).

Subgroup analyses were conducted with efficacy outcomes for carbapenem-resistant Gram-negative bacilli, *K. pneumoniae, P. aeruginosa* and *Acinetobacter spp*..

### Statistical Analysis

We calculated treatment effects as risk ratios (RRs) and 95% confidence interval (CI)s for each outcome with the modified random-effects model (DerSimonian and Laird inverse variance) [21]. For the outcome of day 28 all-cause mortality and nephrotoxicity, RRs <1 favor newer antibiotic. Regarding to the clinical and microbiological response, RRs > 1 favor newer antibiotic groups. The prediction interval was performed in random-effects meta-analysis to predict the possible underlying effect in future trials. The between-study heterogeneity was assessed by using I-squared (*I*^*2*^) statistic, where *I*^*2*^ ≤ 30%, between 30% and 50%, between 50% and 75%, and ≥ 75% were considered to indicate low, moderate, substantial and considerable heterogeneity, respectively [21]. P-values for the *I*^*2*^ statistic were derived from the chi-square test. P-values less than 0.05 were considered statistically significant. A subgroup analysis was also conducted to produce the estimates of efficacy and safety endpoints for each group of pathogens. The GRADE (Grading of Recommendations Assessment, Development, and Evaluation) approach was used to assess the certainty of the evidence for each outcome and described in the summary of findings table [22].

To investigate the impact of modified random-effects method, sensitivity analyses were performed by using standard random-effects model and recalculating the pooled effect estimates. All statistical analyses were performed with R program version 4.4.2, with the function *metabin* [23].

## RESULTS

A total of eight RCTs with 2,881 patients were analyzed. All RCTs were international multicentre trials, funded by pharmaceutical companies. The study sites were in Asia, North and South America, Europe and Australia. The three IMI-REL CHINA, RESTORE-IMI trials 1 and 2 compared imipenem-cilastatin-relebactam with piperacillin-tazobactam and the combination of colistin plus imipenem, respectively [24,25]. Another three trials tested ceftazidime-avibactam, ceftolozane-tazobactam, or cefiderocol in comparison with meropenem [26–28]. The remaining RCTs compared the use of cefiderocol or sulbactam-durlobactam versus the best available therapy of which 61% were colistin-based regimens or colistin [29,30]. The duration of intravenous antibiotic interventions ranged from 7 to 21 days. The mean age of patients was approximately 60. Male participants were the predominant patient population of all trials. The mean APACHE II score was 15 or higher which estimates the mortality of 12-25% among the trial populations. The number of ventilated patients was similar between intervention and comparison groups while the ICU admission of participants was also reported in selected RCTs. The commonest isolated baseline LRT pathogen were *K. pneumoniae, P. aeruginosa*, and *Acinetobacter* spp. (see Supplementary Figure 1 and Table 6).

### GRADE Assessment

At baseline, the quality of evidence derived from included RCTs was assessed as high certainty in all outcomes. Both absolute and relative effects are shown in the summary of evidence findings table (see Supplementary Table 7).

### Effects of interventions

#### Efficacy outcomes

Efficacy of antibiotic treatments in eligible RCTs was evaluated with all-cause mortality at day 28, favorable clinical response and microbiological response at test-of-cure in the relevant population groups. The mean follow-up duration was approximately 28-35 days per participant for post-treatment testing and safety assessment.

##### Day 28 all-cause mortality

Eight RCTs had the complete data for day 28 all-cause mortality in intent-to-treat (ITT), modified ITT (MITT), microbiological modified ITT (mMITT), and microbiological ITT (mITT) population. Pooled effect estimates showed that the newer antibiotic group had no major difference in day-28 all-cause mortality (16.7%) than the generic antibiotic group (17.3%) (RR 0.97, 95% CI 0.72-1.30, *p* 0.81) (Figure 1A). There was limited heterogeneity across included studies (*I*^*2*^ 30%, *p* 0.19). The prediction interval (0.53-1.78) crossed the line of no effect.

**Figure 1.**
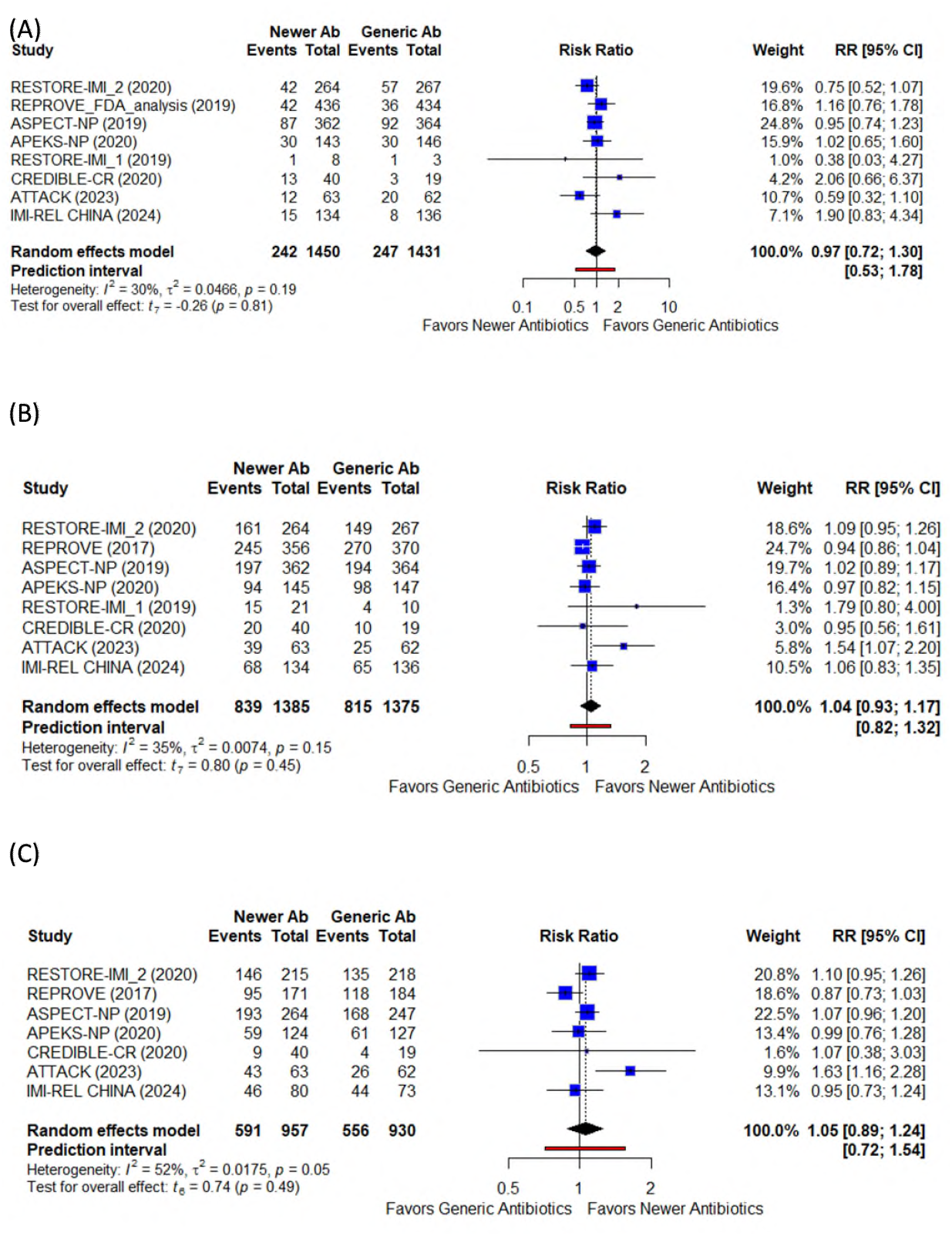
Forest plot of the meta-analysis for efficacy outcomes of newer antibiotics versus generic antibiotics for patients with bacterial HABP/VABP: (A) Day 28 all-cause mortality; (B) clinical response; (C) microbiological response. Forest plot of the meta-analysis for efficacy outcomes of newer antibiotics versus generic antibiotics for patients with bacterial HABP/VABP: (A) Day 28 all-cause mortality in intent-to-treat population (ITT), modified intent-to-treat population (MITT), microbiological modified intent-to-treat population (mMITT) or microbiological intent-to-treat population (mITT); (B) Clinical response in MITT, clinically modified intent-to-treat population (cMITT), ITT, MITT, mMITT or mITT; (C) Microbiological response in mMITT or mITT. RR: risk ratio; CI: confidence interval, I-squared is showing proportion of inconsistency.

##### Favorable clinical response

A favorable clinical response was recorded in 60.6% patients in the newer antibiotic group and 59.3% in the generic treatment group (RR 1.04, 95% CI 0.93-1.17, *p* 0.45) with moderate heterogeneity (*I*^*2*^ 35%, *p* 0.15) (Figure 1B).

##### Favorable microbiological response

A favorable microbiological response was recorded in 61.8% patients in the newer antibiotic group compared to 59.8% in the generic antibiotic group (RR 1.05, 95% CI 0.89-1.24, *p* 0.49) with substantial heterogeneity (*I*^*2*^ 52%, *p* 0.05) (Figure 1C).

### Safety outcomes

#### Nephrotoxicity

Regard to trials in which colistin was the comparator, the relative risk of nephrotoxicity was significantly reduced by 70% in the newer antibiotics group (RR 0.30, 95% CI 0.11-0.79, *p* 0.03). Importantly, there was little heterogeneity between the included trials (*I*^*2*^ 0%, *p* 0.48) (Figure 2).

**Figure 2.**
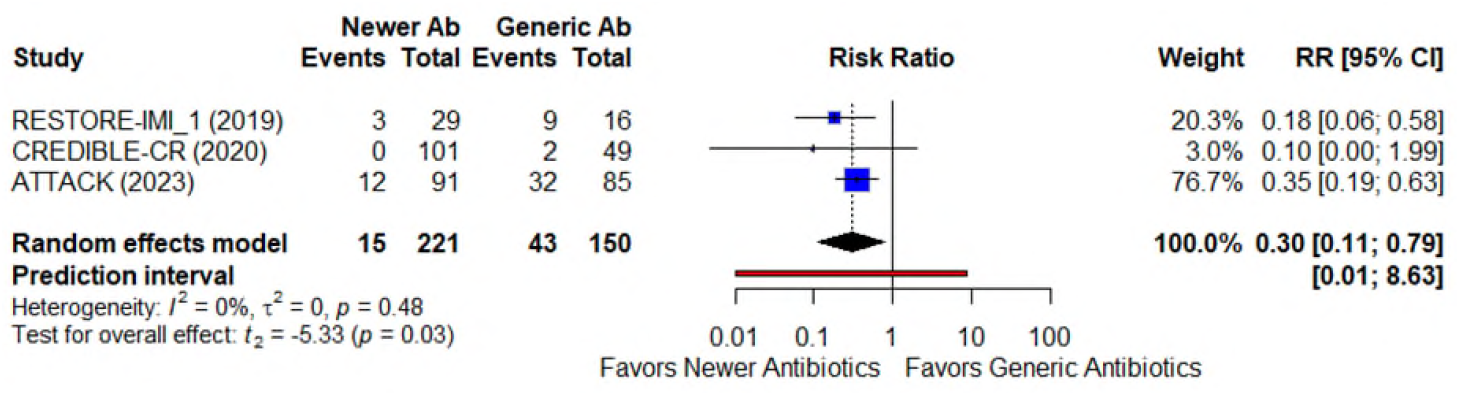
Forest plot of the meta-analysis for safety outcomes of newer antibiotics versus generic antibiotics (Colistin as comparators) for patients with bacterial HABP/VABP: Nephrotoxicity in safety population.

### Sensitivity and subgroup analyses

Pooled estimates of efficacy and safety endpoints showed the similarity in both modified and classic random-effects models (see Supplementary Table 8).

#### Per pathogen Day 28 all-cause mortality

In the subgroup of carbapenem-resistant Gram-negative organisms, 28-day all-cause mortality was 22.0% in those in the newer antibiotic group and 23.5% in the generic antibiotic group (RR 0.91, 95%CI 0.39-2.11). The only organism-specific subgroup in which the pooled effect estimates showed a significant reduction in 28-day all-cause mortality was with *K. pneumoniae* (RR 0.73, 95%CI 0.55-0.99). For HABP/VABP caused by *P. aeruginosa* and *Acinetobacter spp*., the day 28 mortality was similar between the newer antibiotics and the generic antibiotics (RR 1.15, 95%CI 0.09-14.54 and RR 1.05, 95%CI 0.21-5.40). There was moderate heterogeneity across included studies (*I*^*2*^ 32%, *p* 0.13) (Figure 3).

**Figure 3.**
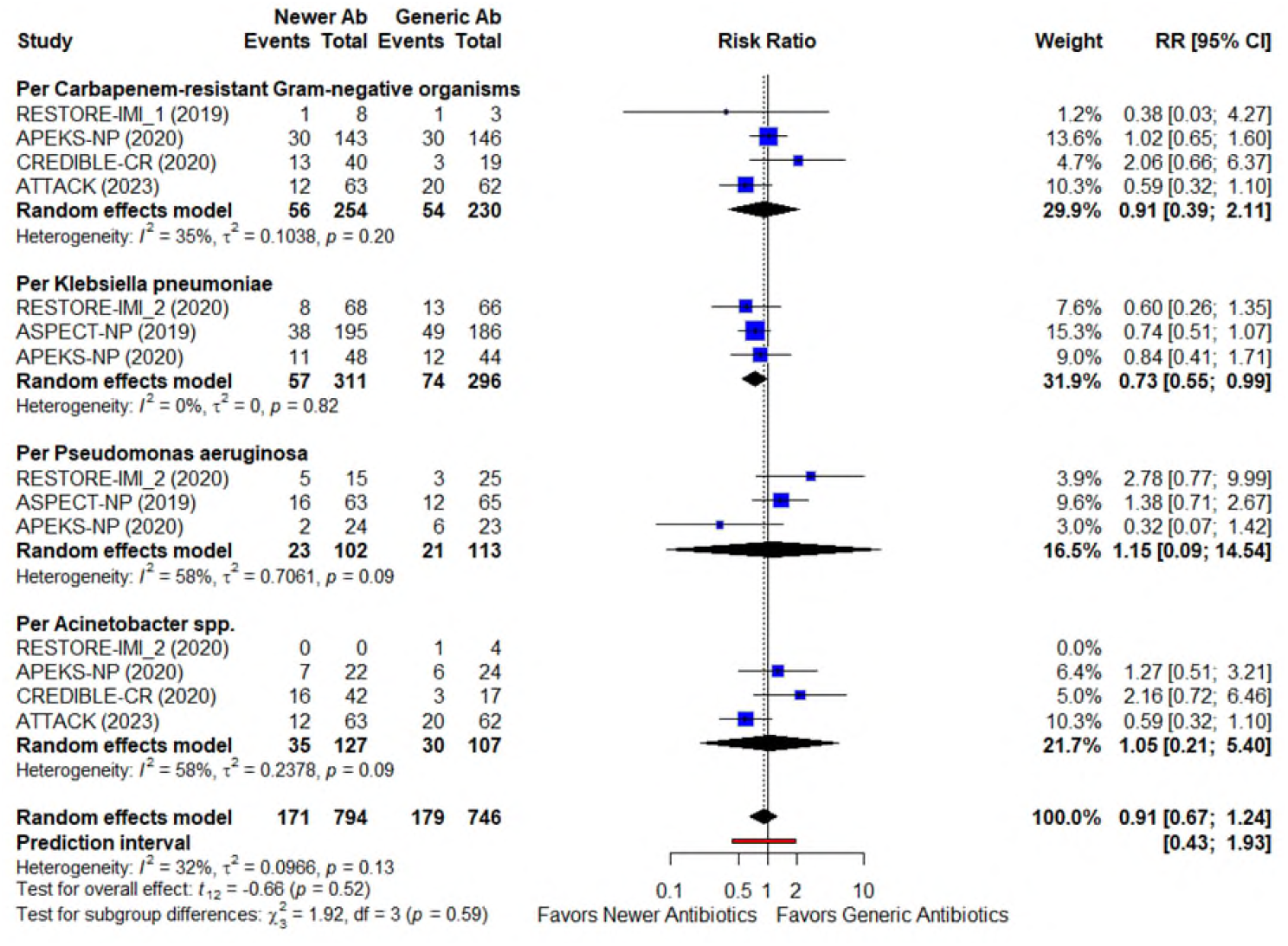
Forest plot of the subgroup analysis for efficacy outcomes of newer antibiotics versus generic antibiotics for patients with bacterial HABP/VABP: Per pathogen Day 28 all-cause mortality.

#### Per pathogen favorable clinical response at test-of-cure

Regarding clinical response, newer antibiotics were not significantly better than generic comparators for bacterial HABP/VABP caused by carbapenem-resistant Gram-negative bacilli, *K. pneumoniae* and *Acinetobacter spp*. (RR 1.21, 95%CI 0.86-1.70; RR 1.05, 95%CI 0.98-1.14 and RR 1.14, 95%CI 0.68-1.89, respectively). In contrast, generic antibiotic treatment was favored over newer therapy in the clinical response of patients with HABP/VABP caused by *P. aeruginosa* (RR 0.88, 95%CI 0.77-1.00). There was little heterogeneity between results from the trials within the pathogen subgroups (*I*^*2*^ 0%) (Figure 4).

**Figure 4.**
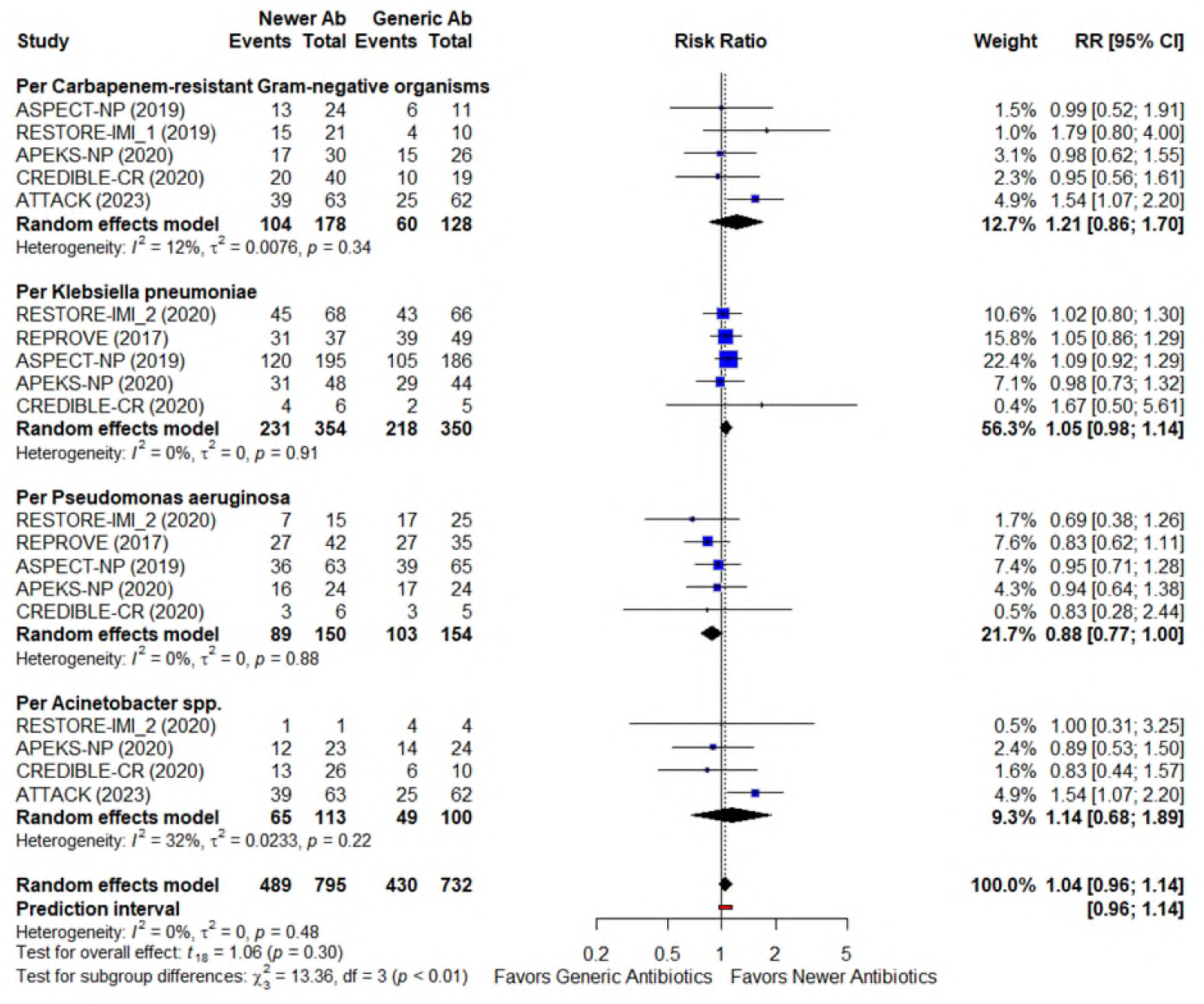
Forest plot of the subgroup analysis for efficacy outcomes of newer antibiotics versus generic antibiotics for patients with bacterial HABP/VABP: Per pathogen Favorable clinical response at test-of-cure.

#### Per pathogen favorable microbiological response at test-of-cure

For the subgroup analysis of microbiological response, newer antibiotic therapy was favored over generic antibiotics (p < 0.01) in the treatment of HABP/VABP caused by carbapenem-resistant Gram-negative bacilli *and Acinetobacter spp*. (RR 1.50, 95%CI 1.18-1.90 and RR 1.44, 95%CI 1.09-1.90). For HABP/VABP caused by *K. pneumoniae* and *P. aeruginosa*, the microbiological response did not differ significantly between the newer antibiotics and the generic antibiotics (RR = 1.10, 95% CI 0.86-1.39 and RR 1.09, 95%CI 0.87-1.36). There was no substantial heterogeneity between results from the trials within the pathogen subgroups (*I*^*2*^ 17%) (Figure 5).

**Figure 5.**
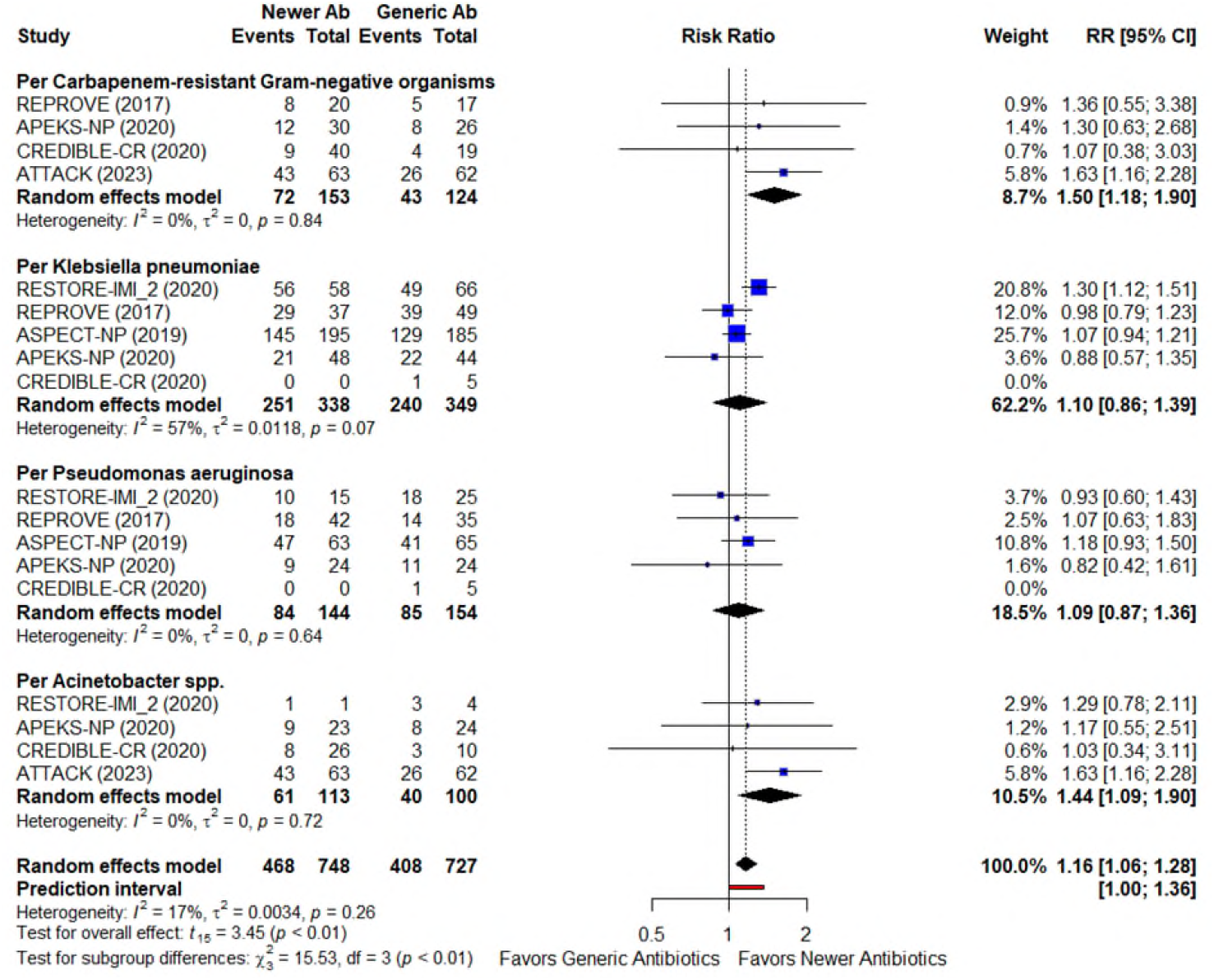
Forest plot of the subgroup analysis for efficacy outcomes of newer antibiotics versus generic antibiotics for patients with bacterial HABP/VABP: Per pathogen Favorable microbiological response at test-of-cure.

## DISCUSSION

In this systematic review and meta-analysis, we showed there were no major improvements using new antibiotics as compared to generic standard of care with respect to 28-day all-cause mortality, favorable clinical response, or favorable microbiological response. Among patients with bacterial HAP/VAP associated with carbapenem-resistant Gram-negative bacilli, the new antibiotic treatments regimes were not associated with a reduction in mortality at day 28 nor in improvements in clinical response compared to generic therapies. Improvement in microbiological response was seen with the new antibiotics for carbapenem-resistant Gram-negative bacilli *and Acinetobacter spp*. compared to generic therapies.

It is noteworthy that for four of the five new antibiotics, their pivotal HABP/VABP trials used for approval by regulatory agencies utilized carbapenem therapy as the comparator. The pooled effect estimates for the key clinical outcomes (all-cause mortality at day 28 and favorable clinical response) favored the new antibiotics and the 95% confidence intervals around the point estimate included a clinically important benefit. However, given that the differentiating features of all of these new antibiotics is activity against carbapenem-resistant Gram-negative organisms, it is surprising that no improvements in mortality, favorable clinical response or favorable microbiological response were observed in this meta-analysis. When the analysis was restricted to patients only with carbapenem resistant organisms, there was no improvement in all-cause mortality nor favorable clinical response with new antibiotics, although an improvement in favorable microbiological response was seen.

One potential explanation for these “negative” results is that the sample size for each of the trials was relatively small (for example, none had a total sample size of more than 1000) and therefore they may have been underpowered to show a significant difference in outcome [31]. This may be because the clinical effect size is quite small despite the *in vitro* attributes of the new antibiotics. Another potential explanation is that VABP is clinically diagnosed but it is not the primary cause of mortality among those trial populations, then the drugs have no effect on patients’ outcomes. It could also be that unexpectedly deleterious effects from some new antibiotics counteract beneficial effects from other new antibiotics. In this regard the most extreme results for all-cause mortality were a relative risk of 2.06 (95%CI 0.66-6.37) for cefiderocol in the CREDIBLE-CR trial compared to 0.59 (95%CI 0.32-1.10) for sulbactam-durlobactam in the ATTACK trial. However, the total weight contributed to the 28-day all-cause mortality meta-analysis by these two trials was just 14.9% suggesting that this explanation is not the most likely one.

The key strength of this meta-analysis is that sub-group analyses have shown a statistically significant favorability of newer antibiotic treatments over generic therapies in reduction in 28 days all-cause mortality for *Klebsiella* infections, and improvement in favorable microbiological response for infections with carbapenem resistant organisms and with *Acinetobacter* infections. Those positive results highlight the need to use newer antibiotics as specific as possible to treat the culprit pathogen rather than targeting at a big group of Gram-negative bacteria or resistant organisms. Furthermore, it must be mentioned that 12 sub-group analyses were performed so there is a risk that “multiplicity” of analyses has contributed to 3 of these sub-group analyses being found to be statistically significant [32]. Although the clinical response in patients with *P aeruginosa* infections receiving generic antibiotics was favorable, it emphasizes that those current standard of care therapies remain their activity against this organism.

Alternatively, it could be that endpoints such as all-cause mortality or favorable clinical response are greatly affected by underlying diseases and less so by the relative effectiveness of the new antibiotics or their generic comparators. By definition, patients with HABP and VABP have been hospitalized due to disease processes sufficiently severe to require hospitalization. This is accentuated when patients have VABP as these patients are all sick enough to require ICU admission. In contrast, favorable microbiological response was more frequently observed in patients with carbapenem resistant organisms or *Acinetobacter*. It could be speculated that microbiological response is more dependent on the in vitro activity of an antibiotic than the underlying diseases of a patient which may explain this difference. Nevertheless, this is not a “patient-centered” outcome measure and therefore has less relevance to the clinical need for a new antibiotic.

This study has several limitations. Firstly, there were only eight included RCTs in this review. Each eligible trial was conducted with the small total sample size which can lead to the lack of major differences in treatment effect between new antibiotics and generic therapies for HABP and VABP. Secondly, only a few studies assessed data on nephrotoxicity, all-cause mortality at day 28, clinical response and microbiological response in carbapenem-resistant Gram-negative bacilli and three Gram-negative pathogens. Thirdly, we have combined all the newer antibiotics into one category and each newer drug was underpowered within single RCT. We did not have enough studies to do subgroup analysis based on the combinations of newer therapies. Finally, those limitations suggested the question if larger studies or adding more studies would push this result to significance.

To our knowledge, this is the first systematic review and meta-analysis of assessment of efficacy and safety between newer and generic antibiotic treatment for bacterial HABP/VABP. The current status, as reflected in many of the trials we have included, is that newer antibiotics are compared to generic antibiotics without in vitro activity against carbapenem-resistant organisms. It is our opinion that new antibiotics which have unique antibacterial attributes (e.g., activity against carbapenem resistant organisms) should be compared in RCTs to standard of care antibiotics for these same carbapenem resistant organisms. This is especially pressing in low- and middle-income countries (especially in Asia) where trial results need to be balanced against the cost of new antibiotics.

## Data Availability

All data in the present work are published and available as detailed in the manuscript.

## Author contributions

Study conception and design: A. N. and D. P. Data extraction and formal analysis: A. N., Y. E., and D. P. Methodology: A. N., Y. E., Y. M., and D. P. Supervision: L. T. Visualization: P. T. D. Writing—original draft: A. N. and D. P. Writing—review and editing: All authors.

## Potential conflicts of interest

No reported conflicts of interest between all authors.

## Notes

### Competing Interest Statement

The authors have declared no competing interest.

### Funding Statement

This study did not receive any funding.

