## Supplementary Materials for "Efficacy and safety of newer antibiotics versus generic antibiotics for hospital-acquired bacterial pneumonia and ventilator-associated bacterial pneumonia: a systematic review and meta-analysis of randomized controlled trials"

### Contents


1. Supplementary Table 1: PRISMA Checklist for reporting systematic review 2
2. Supplementary Table 2: PRISMA 2020 Checklist for abstract reporting in systematic review 5
3. Supplementary Text 1: Search Strategy 6
4. Supplementary Text 2: Eligibility Criteria 11
5. Supplementary Table 3: CONSTORT score of eligible randomized controlled trials 12
6. Supplementary Table 4: Risk of bias assessment 13
7. Supplementary Table 5: Clinical response and microbiological response at the visit 14
8. Supplementary Figure 1: PRISMA 2020 flow diagram of the retained randomized controlled trials 16
9. Supplementary Table 6: Characteristics of eligible randomized controlled trials 18
10. Supplementary Table 7: Summary of evidence findings 20
11. Supplementary Table 8: Sensitivity analysis 21

**Supplementary Table 1.** PRISMA 2020 checklist for reporting systematic review.

### PRISMA 2020 Checklist

| Section and Topic | Item # | Checklist item | Location where item is reported |
|---|---|---|---|
| **TITLE** | | | |
| Title | 1 | Identify the report as a systematic review. | 1 |
| **ABSTRACT** | | | |
| Abstract | 2 | See the PRISMA 2020 for Abstracts checklist. | 1 |
| **INTRODUCTION** | | | |
| Rationale | 3 | Describe the rationale for the review in the context of existing knowledge. | 3 |
| Objectives | 4 | Provide an explicit statement of the objective(s) or question(s) the review addresses. | 3 |
| **METHODS** | | | |
| Eligibility criteria | 5 | Specify the inclusion and exclusion criteria for the review and how studies were grouped for the syntheses. | Supplementary Text 2 |
| Information sources | 6 | Specify all databases, registers, websites, organisations, reference lists and other sources searched or consulted to identify studies. Specify the date when each source was last searched or consulted. | 3-4 |
| Search strategy | 7 | Present the full search strategies for all databases, registers and websites, including any filters and limits used. | Supplementary Text 1 |
| Selection process | 8 | Specify the methods used to decide whether a study met the inclusion criteria of the review, including how many reviewers screened each record and each report retrieved, whether they worked independently, and if applicable, details of automation tools used in the process. | 4 |
| Data collection process | 9 | Specify the methods used to collect data from reports, including how many reviewers collected data from each report, whether they worked independently, any processes for obtaining or confirming data from study investigators, and if applicable, details of automation tools used in the process. | 4 |
| Data items | 10a | List and define all outcomes for which data were sought. Specify whether all results that were compatible with each outcome domain in each study were sought (e.g. for all measures, time points, analyses), and if not, the methods used to decide which results to collect. | 4-5 |
| | 10b | List and define all other variables for which data were sought (e.g. participant and intervention characteristics, funding sources). Describe any assumptions made about any missing or unclear information. | 4-5 |
| Study risk of bias assessment | 11 | Specify the methods used to assess risk of bias in the included studies, including details of the tool(s) used, how many reviewers assessed each study and whether they worked independently, and if applicable, details of automation tools used in the process. | 4 |
| Effect measures | 12 | Specify for each outcome the effect measure(s) (e.g. risk ratio, mean difference) used in the synthesis or presentation of results. | 5 |
| Synthesis methods | 13a | Describe the processes used to decide which studies were eligible for each synthesis (e.g. tabulating the study intervention characteristics and comparing against the planned groups for each synthesis (item #5)). | 5 |
| | 13b | Describe any methods required to prepare the data for presentation or synthesis, such as handling of missing summary statistics, or data conversions. | 5 |
| | 13c | Describe any methods used to tabulate or visually display results of individual studies and syntheses. | 5 |
| | 13d | Describe any methods used to synthesize results and provide a rationale for the choice(s). If meta-analysis was performed, describe the model(s), method(s) to identify the presence and extent of statistical heterogeneity, and software package(s) used. | 5 |
| | 13e | Describe any methods used to explore possible causes of heterogeneity among study results (e.g. subgroup analysis, meta-regression). | 5 |

**Supplementary Table 1.** PRISMA 2020 checklist for reporting systematic review.

### PRISMA 2020 Checklist

| Section and Topic | Item # | Checklist item | Location where item is reported |
|---|---|---|---|
| | 13f | Describe any sensitivity analyses conducted to assess robustness of the synthesized results. | 5 |
| Reporting bias assessment | 14 | Describe any methods used to assess risk of bias due to missing results in a synthesis (arising from reporting biases). | 4 |
| Certainty assessment | 15 | Describe any methods used to assess certainty (or confidence) in the body of evidence for an outcome. | 9 |
| **RESULTS** | | | |
| Study selection | 16a | Describe the results of the search and selection process, from the number of records identified in the search to the number of studies included in the review, ideally using a flow diagram. | Supplementary Figure 1 |
| | 16b | Cite studies that might appear to meet the inclusion criteria, but which were excluded, and explain why they were excluded. | Supplementary Figure 1 |
| Study characteristics | 17 | Cite each included study and present its characteristics. | Supplementary Table 7 |
| Risk of bias in studies | 18 | Present assessments of risk of bias for each included study. | Supplementary Table 4 |
| Results of individual studies | 19 | For all outcomes, present, for each study: (a) summary statistics for each group (where appropriate) and (b) an effect estimate and its precision (e.g. confidence/credible interval), ideally using structured tables or plots. | 7-12 |
| Results of syntheses | 20a | For each synthesis, briefly summarise the characteristics and risk of bias among contributing studies. | 7-12 |
| | 20b | Present results of all statistical syntheses conducted. If meta-analysis was done, present for each the summary estimate and its precision (e.g. confidence/credible interval) and measures of statistical heterogeneity. If comparing groups, describe the direction of the effect. | 7-12 |
| | 20c | Present results of all investigations of possible causes of heterogeneity among study results. | 7-12 |
| | 20d | Present results of all sensitivity analyses conducted to assess the robustness of the synthesized results. | 7-12 |
| Reporting biases | 21 | Present assessments of risk of bias due to missing results (arising from reporting biases) for each synthesis assessed. | Supplementary Table 4 |
| Certainty of evidence | 22 | Present assessments of certainty (or confidence) in the body of evidence for each outcome assessed. | Supplementary Table 8 |
| **DISCUSSION** | | | |
| Discussion | 23a | Provide a general interpretation of the results in the context of other evidence. | 13 |
| | 23b | Discuss any limitations of the evidence included in the review. | 14 |
| | 23c | Discuss any limitations of the review processes used. | 13 |
| | 23d | Discuss implications of the results for practice, policy, and future research. | 14 |
| **OTHER INFORMATION** | | | |
| Registration and protocol | 24a | Provide registration information for the review, including register name and registration number, or state that the review was not registered. | 3 |
| | 24b | Indicate where the review protocol can be accessed, or state that a protocol was not prepared. | 3 |

**Supplementary Table 1.** PRISMA 2020 checklist for reporting systematic review.

### PRISMA 2020 Checklist

| Section and Topic | Item # | Checklist item | Location where item is reported |
|---|---|---|---|
| | 24c | Describe and explain any amendments to information provided at registration or in the protocol. | Not applicable |
| Support | 25 | Describe sources of financial or non-financial support for the review, and the role of the funders or sponsors in the review. | Protocol |
| Competing interests | 26 | Declare any competing interests of review authors. | Protocol |
| Availability of data, code and other materials | 27 | Report which of the following are publicly available and where they can be found: template data collection forms; data extracted from included studies; data used for all analyses; analytic code; any other materials used in the review. | Protocol |

*From:* Page MJ, McKenzie JE, Bossuyt PM, Boutron I, Hoffmann TC, Mulrow CD, et al. The PRISMA 2020 statement: an updated guideline for reporting systematic reviews. BMJ 2021;372:n71. doi: 10.1136/bmj.n71

For more information, visit: http://www.prisma-statement.org/

**Supplementary Table 2** PRISMA 2020 Checklist for abstract reporting in systematic review

### PRISMA 2020 for Abstracts Checklist

| Section and Topic | Item # | Checklist item | Reported (Yes/No) |
|---|---|---|---|
| **TITLE** | | | |
| Title | 1 | Identify the report as a systematic review. | Yes |
| **BACKGROUND** | | | |
| Objectives | 2 | Provide an explicit statement of the main objective(s) or question(s) the review addresses. | Yes |
| **METHODS** | | | |
| Eligibility criteria | 3 | Specify the inclusion and exclusion criteria for the review. | Yes |
| Information sources | 4 | Specify the information sources (e.g. databases, registers) used to identify studies and the date when each was last searched. | Yes |
| Risk of bias | 5 | Specify the methods used to assess risk of bias in the included studies. | No |
| Synthesis of results | 6 | Specify the methods used to present and synthesise results. | Yes |
| **RESULTS** | | | |
| Included studies | 7 | Give the total number of included studies and participants and summarise relevant characteristics of studies. | Yes |
| Synthesis of results | 8 | Present results for main outcomes, preferably indicating the number of included studies and participants for each. If meta-analysis was done, report the summary estimate and confidence/credible interval. If comparing groups, indicate the direction of the effect (i.e. which group is favoured). | Yes |
| **DISCUSSION** | | | |
| Limitations of evidence | 9 | Provide a brief summary of the limitations of the evidence included in the review (e.g. study risk of bias, inconsistency and imprecision). | No |
| Interpretation | 10 | Provide a general interpretation of the results and important implications. | Yes |
| **OTHER** | | | |
| Funding | 11 | Specify the primary source of funding for the review. | No |
| Registration | 12 | Provide the register name and registration number. | No |

*From:* Page MJ, McKenzie JE, Bossuyt PM, Boutron I, Hoffmann TC, Mulrow CD, et al. The PRISMA 2020 statement: an updated guideline for reporting systematic reviews. BMJ 2021;372:n71. doi: 10.1136/bmj.n71

**Supplementary Text 1.** Search strategy

**Database: PubMed, search date, 31 July 2023**
**Newer antibiotics vs Carbapenem**
**65 results**
**Search:**
((((((("Adult"[MeSH Terms] OR "Young Adult"[MeSH Terms]) AND ("Healthcare-Associated Pneumonia"[MeSH Terms] OR "pneumonia, ventilator associated"[MeSH Terms])) OR ("Critical Care"[MeSH Terms] OR "Intensive Care Units"[MeSH Terms])) AND ("avibactam ceftazidime drug combination"[Supplementary Concept] OR "ceftazidime monobactam"[Supplementary Concept])) OR "ceftolozane tazobactam drug combination"[Supplementary Concept] OR "cefiderocol"[Supplementary Concept] OR "sulbactam-durlobactam"[Supplementary Concept] OR "delafloxacin"[Supplementary Concept]) AND ("Meropenem"[MeSH Terms]) OR ("Imipenem"[MeSH Terms] OR "cilastatin, imipenem drug combination"[MeSH Terms]) OR ("Ertapenem"[MeSH Terms] OR "Doripenem"[MeSH Terms])) AND "Randomized Controlled Trial"[Publication Type]) AND ((fft[Filter]) AND (2013:2023[pdat]))

**PubMed, search date, 07 September 2023**
**Newer antibiotics vs other antibiotic groups**
**12 results**
**Search:**
(((("Healthcare-Associated Pneumonia"[MeSH Terms] OR "pneumonia, ventilator associated"[MeSH Terms]) OR ("Critical Care"[MeSH Terms] OR "Intensive Care Units"[MeSH Terms])) AND (("avibactam ceftazidime drug combination"[Supplementary Concept] OR "ceftazidime monobactam"[Supplementary Concept]) OR ("ceftolozane tazobactam drug combination"[Supplementary Concept]) OR ("cefiderocol"[Supplementary Concept]) OR ("sulbactam-durlobactam"[Supplementary Concept]) OR ("delafloxacin"[Supplementary Concept]))) AND (("Piperacillin, Tazobactam Drug Combination"[Mesh]) OR ("Colistin"[Mesh]) OR ("Polymyxin B"[Mesh]) OR ("Cephalosporins"[Mesh]))) AND (("Randomized Controlled Trial"[Publication Type]))

**PubMed, search date, 07 January 2025**
**Imipenem/cilastatin/relebactam**
**90 results**
**Search:**
imipenem/cilastatin/relebactam AND (2013:2025[pdat])

**Database: CENTRAL, search date, 25 July 2023**
https://www.cochranelibrary.com/central/about-central
https://www.cochranelibrary.com/advanced-search
https://www.cochranelibrary.com/advanced-search/search-manager
**Newer antibiotics vs Carbapenem**
**19 results**
**Search:**
Search Name: VAP: Newer antibiotics vs carbapenems (Cochrane)
Date Run: 25/07/2023 00:35:21

| ID | Search Hits |
| --- | --- |
| #1 | MeSH descriptor: [Pneumonia, Ventilator-Associated] explode all trees |
| #2 | MeSH descriptor: [Adult] explode all trees |
| #3 | MeSH descriptor: [Intensive Care Units] explode all trees |
| #4 | (ventilator-associated NEXT pneumonia*) |
| #5 | #1 OR #4 |
| #6 | #1 OR #4 AND #2 |
| #7 | #1 OR #4 AND #2 AND #3 |
| #8 | (ceftazidime NEXT avibactam*) OR (avibactam NEXT ceftazidime*) OR (ceftazidime NEXT monobactam*) OR (monobactam NEXT ceftazidime*) OR (ceftazidime*) |
| #9 | (ceftolozane NEXT tazobactam*) OR (tazobactam NEXT ceftolozane*) OR (ceftolozane*) |
| #10 | (ceftazidime NEXT avibactam*) OR (avibactam NEXT ceftazidime*) OR (ceftazidime NEXT monobactam*) OR (monobactam NEXT ceftazidime*) |
| #11 | MeSH descriptor: [Aztreonam] explode all trees |
| #12 | (cefiderocol*) |
| #13 | (sulbactam NEXT durlobactam*) OR (durlobactam NEXT sulbactam*) |
| #14 | (delafloxacin*) |
| #15 | #8 OR #9 OR #10 OR #11 OR #12 OR #13 OR #14 |
| #16 | MeSH descriptor: [Carbapenems] explode all trees |
| #17 | (meropenem*) OR (imipenem*) OR (imipenem NEXT cilastatin*) OR (cilastatin NEXT imipenem*) OR (ertapenem*) OR (doripenem*) |
| #18 | #16 OR #17 |
| #19 | (Randomized NEXT Controlled NEXT Trial*) OR (Randomized* NEXT Controlled NEXT Trial) OR (Randomized NEXT Controlled* NEXT Trial) OR (RCT*) |
| **#20** | **#5 AND #15 AND #18 AND #19** |

**Search Diary**
**#5 AND #15 AND #18 AND #19**

1. No need to add “adult” in the search strategy as population in those articles is adult.
2. Article 2010: nebulized colistimethate sodium
3. Article 2008: probiotic
4. Article 2000: diagnostic techniques
5. Article 2008: tracheobronchitis

**CENTRAL, search date, 06 September 2023**
**16 results**
**Search:**

Search Name: **Newer antibiotics vs other antibiotic groups**
Date Run: 06/09/2023 18:37:08
Comment:

ID Search Hits
#1 ventilator-associated NEXT pneumonia* 1779
#2 (ceftazidime NEXT avibactam*) OR (avibactam NEXT ceftazidime*) OR (ceftazidime NEXT monobactam*) OR (monobactam NEXT ceftazidime*) OR (ceftazidime*) 1200
#3 (ceftolozane NEXT tazobactam*) OR (tazobactam NEXT ceftolozane*) OR (ceftolozane*) 76
#4 Aztreonam 488
#5 (cefiderocol*) 35
#6 (sulbactam NEXT durlobactam*) OR (durlobactam NEXT sulbactam*) 10
#7 (delafloxacin*) 56
#8 #2 OR #3 OR #4 OR #5 OR #6 OR #7 1741
#9 piperacillin NEXT tazobactam 612
#10 colistin* 590
#11 polymyxin B* 522
#12 cephalosporin* 2989
#13 #9 OR #10 OR #11 OR #12 4511
#14 (Randomized NEXT Controlled NEXT Trial*) OR (Randomized* NEXT Controlled NEXT Trial) OR (Randomized NEXT Controlled* NEXT Trial) OR (RCT*) 691650
#15 #1 AND #8 AND #13 AND #14 **16**

**Database: Scopus, search date, 26 July 2023**
https://www.scopus.com
**Newer antibiotics vs Carbapenem**
**16 results**
**Search:**
( TITLE-ABS-KEY ( ventilator-associated AND pneumonia* ) ) AND ( TITLE-ABS-KEY ( ceftazidime-avibactam OR ceftazidime-avibactam* OR aztreonam OR aztreonam* OR ceftolozane-tazobactam OR ceftolozane-tazobactam* OR cefiderocol OR cefiderocol* OR sulbactam-durlobactam OR sulbactam-durlobactam* OR delafloxacin OR delafloxacin* ) ) AND ( TITLE-ABS-KEY ( carbapenem OR carbapenem* OR meropenem OR meropenem* OR imipenem OR imipenem* OR imipenem-cilastatin OR imipenem-cilastatin* OR ertapenem OR ertapenem* OR doripenem OR doripenem* ) ) AND ( TITLE-ABS-KEY ( randomise* AND control* AND trial* OR rct* ) )
**(Limited from 2013 to 2023)**

**Scopus, search date, 11 September 2023**
**Newer antibiotics vs other antibiotic groups**
**8 results**
**Search:**
( TITLE-ABS-KEY ( ventilator-associated AND pneumonia* ) ) AND ( ( TITLE-ABS-KEY ( ceftazidime-avibactam OR ceftazidime-avibactam* OR aztreonam OR aztreonam* OR ceftolozane-tazobactam OR ceftolozane-tazobactam* OR cefiderocol OR cefiderocol* OR sulbactam-durlobactam OR sulbactam-durlobactam* OR delafloxacin OR delafloxacin* ) ) ) AND ( ( TITLE-ABS-KEY ( piperacillin-tazobactam* ) ) OR ( TITLE-ABS-KEY ( colistin* ) ) OR ( TITLE-ABS-KEY ( polymyxin AND b* ) ) OR ( TITLE-ABS-KEY ( cephalosporin* ) ) ) AND ( TITLE-ABS-KEY ( randomise* AND control* AND trial* OR rct* ) ) AND PUBYEAR > 2012 AND PUBYEAR < 2024

**Database: Ovid MEDLINE, search date, 28 July 2023**
[https://ovidsp.ovid.com](https://ovidsp.ovid.com)
Ovid: Search Form
**Newer antibiotics vs Carbapenem**
**125 results**
**Search:**
(ventilator-associated pneumonia or ventilator-associated pneumonia*) AND (ceftazidime-avibactam or ceftazidime-avibactam* or aztreonam or aztreonam* or ceftolozane-tazobactam or ceftolozane-tazobactam* or cefiderocol or cefiderocol* or sulbactam-durlobactam or sulbactam-durlobactam* or delafloxacin or delafloxacin*) AND (carbapenem or carbapenem* or meropenem or meropenem* or imipenem or imipenem* or imipenem-cilastatin or imipenem-cilastatin* or ertapenem or ertapenem* or doripenem or doripenem*) AND ((randomise* and control* and trial*) or rct*)

**Ovid MEDLINE, search date, 11 September 2023**
**Newer antibiotics vs other antibiotic groups**
**95 results**
**Search:**
(ventilator-associated pneumonia or ventilator-associated pneumonia*) AND (ceftazidime-avibactam or ceftazidime-avibactam* or aztreonam or aztreonam* or ceftolozane-tazobactam or ceftolozane-tazobactam* or cefiderocol or cefiderocol* or sulbactam-durlobactam or sulbactam-durlobactam* or delafloxacin or delafloxacin*) AND (piperacillin-tazobactam* or colistin* or polymyxin* or cephalosporin*) AND ((randomise* and control* and trial*) or rct*) limited from 2013 to 2023

**Database: Clinical Trials, search date, 14 August 2023**
https://clinicaltrials.gov
**Newer antibiotics vs Carbapenem**
**122 results**
**Search:**
Condition or disease: ventilator-associated pneumonia
Intervention/Treatment: antibiotics
Age: 18 or older

**Database: Google Scholar, search date, 14 Aug 2023**
**Newer antibiotics vs Carbapenem**
**150 results**
**Search:**
allintitle: ventilator associated pneumonia "ceftazidime avibactam" OR aztreonam OR "ceftolozane tazobactam" OR cefiderocol OR "sulbactam durlobactam" OR delafloxacin OR carbapenem OR meropenem OR imipenem OR "imipenem cilastatin" OR ertapenem OR doripenem

**Advanced search**
With all of the words: ventilator associated pneumonia
With the exact phrase: randomised controlled trial
With at least one of the words: "ceftazidime avibactam" aztreonam "ceftolozane tazobactam" cefiderocol "sulbactam durlobactam" delafloxacin carbapenem meropenem imipenem "imipenem cilastatin" ertapenem doripenem
Without the words: NA
Where my words occur: in the title of the article
Return articles dated between: 2013 -2023
Include patents and citations

**Supplementary Text 2.** Eligibility criteria

**Inclusion Criteria:** The search terms were developed according to the PICOS criteria as follow:

- **Participants (P)**: adult participants (aged 18 years and older) with hospital-acquired bacterial pneumonia and ventilator-associated bacterial pneumonia.
- **Intervention (I)**: newer antibiotics (approved by the USA Food and Drug Administration for hospital-acquired bacterial pneumonia and ventilator-associated bacterial pneumonia, from 2013 to 2023), including Ceftazidime-avibactam, Cefiderocol, Imipenem-cilastatin-relabactam, Sulbactam-durlobactam and Ceftolozane-tazobactam.
- **Comparison (C)**: generic antibiotics, including Carbapenems, Piperacillin-tazobactam, Colistin, Polymyxin B and third and fourth generation Cephalosporins.
- **Outcomes (O)**: efficacy and safety measures, including day 28 all-cause mortality, clinical response, microbiological response and nephrotoxicity.
- **Study type (S)**: randomized controlled trials.

**Exclusion Criteria**

- Duplicate studies
- Review papers
- Paediatric studies
- Observational studies
- Studies included placebo controls
- Study with incomplete data
- Study population including

  -community-acquired pneumonia,
  -healthy volunteers.

**Supplementary Table 3.** CONSORT score of eligible randomized controlled trials

| Study ID | Study | CONSORT score |
|---|---|---|
| 1 | **RESTORE-IMI_2** | 25/25 |
| 2 | **REPROVE** | 24/25 |
| 3 | **ASPECT-NP** | 24/25 |
| 4 | **APEKS-NP** | 24/25 |
| 5 | **RESTORE-IMI_1** | 21/25 |
| 6 | **CREDIBLE-CR** | 24/25 |
| 7 | **ATTACK** | 23/25 |
| 8 | **IMI-REL CHINA** | 23/25 |

**Supplementary Table 4**. Risk of bias assessment

| Study | D1 | D2 | D3 | D4 | D5 | Overall |
|---|---|---|---|---|---|---|
| **RESTORE-IMI 2 (2020)** | | | | | | |
| **REPROVE (2017)** | + | + | + | + | + | + |
| **ASPECT-NP (2019)** | | | | | | |
| **APEKS-NP (2020)** | + | + | + | + | + | + |
| **RESTORE-IMI 1 (2019)** | | | | | | |
| **CREDIBLE-CR (2020)** | + | ! | + | + | + | ! |
| **ATTACK (2023)** | | | | | | |
| **IMI-REL CHINA (2024)** | + | + | + | + | + | + |

Domains:

D1: Bias arising from the randomization process

D2: Bias due to deviations from intended interventions

D3: Bias due to missing outcome data

D4: Bias in measurement of the outcome

D5: Bias in selection of the reported result

Judgement:

+ Low risk

! Some concern

High risk

**Supplementary Table 5**. Clinical response and microbiological response at the visit: (A) clinical response (CR); (B): microbiological response (MR). (A)

| Study | Clinical response | Definition of CR | Time point | Definition of time point |
|---|---|---|---|---|
| **RESTORE-IMI 2** | Favorable clinical response | • Resolution of baseline HAP/VAP signs/symptoms, AND Improve chest X-Ray; OR<br>• No additional HAP/VAP signs/symptoms, AND no non-study antibacterial therapy for HAP/VAP at the visit. | At EFU visit | 7-14 days post-EOT |
| **REPROVE** | Clinical cure | | At TOC | 21-25 days after randomization |
| **ASPECT-NP** | Clinical cure | | At TOC | 7-14 days post-EOT |
| **APEKS-NP** | Clinical cure | | At TOC | 7 days +/- 2 days post-EOT |
| **RESTORE-IMI 1** | Favorable clinical response | | Day 28 | 28 days after randomization |
| **CREDIBLE-CR** | Clinical cure | | At TOC | 7 days +/- 2 days post-EOT |
| **ATTACK** | Clinical cure | | At TOC | 7 days +/- 2 days post-EOT |
| **IMI-REL CHINA** | Favorable clinical response | Supplementary Table 3 (pre-proof article) | At EFU and at EOT | EFU: 7-14 days post-EOT |

EFU = Early follow-up, EOT = End of therapy, TOC = Test-of-cure.

(B)

| Study | Microbiological response | Definition of MR | Time point | Definition of time point |
|---|---|---|---|---|
| **RESTORE-IMI 2** | Favorable microbiological response | = eradication + presumed eradication - eradication: an LRT culture at EFU shows eradication of the pathogen found at study entry. -presumed eradication: no culture at EFU AND the patient was assessed as a clinical cure. | At EFU visit | 7-14 days post-EOT |
| **REPROVE** | Favorable microbiological response | = eradication + presumed eradication | At TOC | 21-25 days after randomization |
| **ASPECT-NP** | Microbiological eradication | LRT culture at TOC: $\leq 10^4$ CFU/mL AND reduction $\geq$ 1-log of baseline pathogen. | At TOC | 7-14 days post-EOT |
| **APEKS-NP** | Microbiological eradication | Absence of baseline pathogen. | At TOC | 7 days $\pm$ 2 days post-EOT |
| **RESTORE-IMI 1** | Microbiological eradication | Reduction $\geq$ 1-log of baseline pathogen (e.g., $\geq 10^5$ to $< 10^4$) | 5-9 days post-EOT | |
| **CREDIBLE-CR** | Microbiological eradication | Absence of baseline pathogen. | At TOC | 7 days $\pm$ 2 days post-EOT |
| **ATTACK** | Microbiological response | = eradication + presumed eradication | At TOC | 7 days $\pm$ 2 days post-EOT |
| **IMI-REL CHINA** | Favorable microbiological response | No definition in the pre-proof article and the supplementary | At EFU and at EOT | EFU: 7-14 days post-EOT |

EFU = Early follow-up, EOT = End of therapy, LRT = Low respiratory tract, TOC = Test-of-cure.

**Supplementary Figure 1.** PRISMA 2020 flow diagram of the retained randomized controlled trials

**PRISMA 2020 flow diagram for new systematic reviews which included searches of databases and registers only**

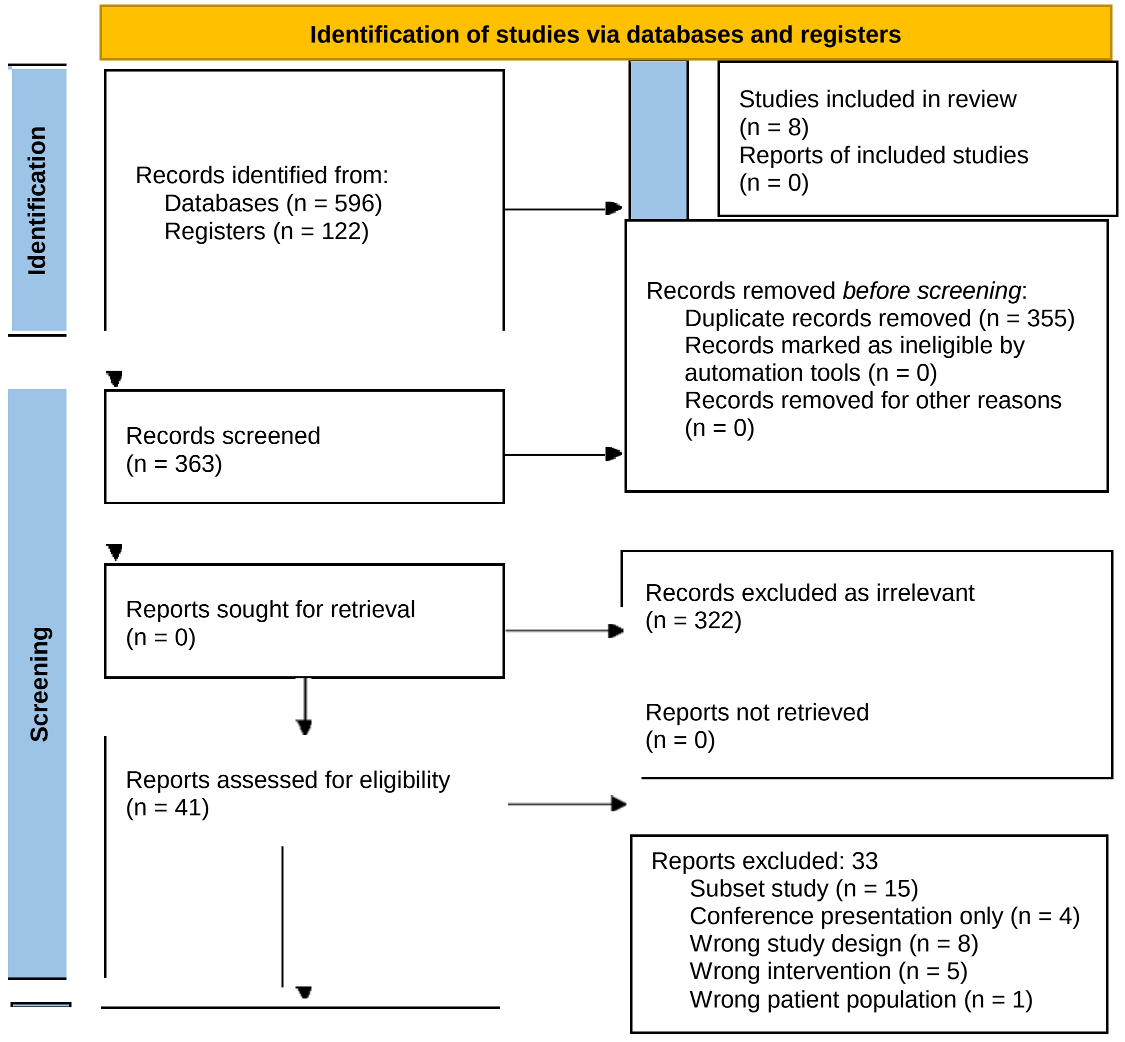


*From:* Page MJ, McKenzie JE, Bossuyt PM, Boutron I, Hoffmann TC, Mulrow CD, et al. The PRISMA 2020 statement: an updated guideline for reporting systematic reviews. BMJ 2021;372:n71. doi: 10.1136/bmj.n71

For more information, visit: http://www.prisma-statement.org/

**PRISMA 2020 flow diagram for new systematic reviews which included searches of databases and registers only**

**Exclusion**

**Wrong study design**

| Article | Wrong study design |
|---|---|
| Cefiderocol as Adjunctive Treatment of Necrotizing Ventilator-associated Pneumonia Due to Extensively Drug-Resistant Pseudomonas aeruginosa | Case report |
| Evaluation of plazomicin, tigecycline, and meropenem pharmacodynamic exposure against carbapenem-resistant Enterobacteriaceae in patients with bloodstream infection or hospital-acquired/ventilator-associated pneumonia from the CARE study (ACHN-490-007) | No RCT |
| Cost effectiveness of ceftolozane/tazobactam compared with meropenem for the treatment of patients with ventilated hospital-acquired bacterial pneumonia and ventilator-associated bacterial pneumonia. | No RCT |
| PIN1 SYSTEMATIC REVIEW AND NETWORK META-ANALYSIS COMPARING THE EFFICACY AND SAFETY OF CEFTOLOZANE/TAZOBACTAM AND COMPARATORS IN THE TREATMENT OF HOSPITAL-ACQUIRED BACTERIAL PNEUMONIA AND VENTILATOR-ASSOCIATED BACTERIAL PNEUMONIA | No RCT |
| Ceftolozane/tazobactam versus colistin in the treatment of ventilator-associated pneumonia due to extensively drug-resistant Pseudomonas aeruginosa | Observational study |
| Carbapenem-resistant Klebsiella pneumoniae among patients with ventilator-associated pneumonia: Evaluation of antibiotic combinations and susceptibility to new antibiotics | In vitro study |
| Efficacy of cefiderocol-vs colistin-containing regimen for treatment of bacteraemic ventilator-associated pneumonia caused by carbapenem-resistant Acinetobacter baumannii in patients with COVID-19 | Observational study |
| Meta-analysis of Clinical Outcomes Using Ceftazidime/Avibactam, Ceftolozane/Tazobactam, and Meropenem/Vaborbactam for the Treatment of Multidrug-Resistant Gram-Negative Infections. | No RCT |

COVID-19 = Coronavirus disease 2019, RCT = Randomized controlled trial.

**Supplementary Table 6.** Characteristics of eligible randomized controlled trials.

| Name of RCTs | Author (Year) | Population | Total patients (Population) | Newer Antibiotics (n) | Generic Antibiotics (n) | Age N/G | Gender (F vs M) N/G | Duration of antibiotics (days) | Ventilated pneumonia N/G | ICU admission N/G | Normal renal function N/G | APACHE II score (mean ± SD) N/G | Baseline LRT pathogen |
|---|---|---|---|---|---|---|---|---|---|---|---|---|---|
| **RESTORE -IMI 2** | Titov et al. (2020) | Aged ≥18 years with HABP or VABP | 531 (MITT) | Imipenem-cilastatin-relebactam (264) | Piperacillin-tazobactam (267) | 60.5 ± 16.9/ 58.8 ± 18.4 | 86 vs 178/ 78 vs 189 | 7-14 | 122/136 | 175/176 | 103/85 | 14.6 ± 6.2/ 14.8 ± 6.7 | *K. pneumoniae , P. aeruginosa, A. calcoaceticus-baumannii complex* |
| **REPROVE** | Torres et al. (2017) | Aged 18-90 years with HAP or VAP | 726 (cMITT) | Ceftazidime - avibactam (356) | Meropenem (370) | 62.1 ± 16.6/ 61.9 ± 17.4 | 88 vs 268/ 96 vs 274 | 7-14 | 154/159 | NA | 286/292 | 14.5 ± 4.01/ 14.9 ± 4.05 | *K. pneumoniae , P. aeruginosa* |
| **ASPECT-NP** | Kollef et al. (2019) | Aged ≥18 years with VAP or ventilated HAP | 726 (ITT) | Ceftolozane-tazobactam (362) | Meropenem (364) | 60.5 ± 16.7/ 59.5 ± 17.2 | 100 vs 262/ 109 vs 255 | 8-14 | The participant s were all ventilated | 334/334 | 294/300 | 17.5 ± 5.2/ 17.4 ± 5.7 | *K. pneumoniae , E. coli, P. aeruginosa* |
| **APEKS-NP** | Wunderink et al. (2020) | Aged ≥18 years with HAP or VAP or HCAP | 292 (MITT) | Cefiderocol (145) | Meropenem (147) | 64.6 ± 14.6/ 65.4 ± 15.1 | 46 vs 99/ 46 vs 101 | 7-14 | 89/86 | 102/97 | 55/61 | 16.0 ± 6.1/ 16.4 ± 6.9 | *K. pneumoniae , P. aeruginosa, A. baumannii* |
| **RESTORE -IMI 1** | Motsch et al. (2019) | Aged ≥18 years with HABP or VABP | 31 (mMITT) | Imipenem-cilastatin-relebactam (21) | Colistin + Imipenem-cilastatin (10) | 59 (19-75)/ 61 (49-80) | 8 vs 13/ 3 vs 7 | 7-21 | 7/2 | NA | 16/7 | 16 (0-26)/ 22 (14-23) | *P. aeruginosa, Klesiella spp,* |

**Supplementary Table 6.** Characteristics of eligible randomized controlled trials.

| | | | | | | | | | | | | | |
|---|---|---|---|---|---|---|---|---|---|---|---|---|---|
| **CREDIBLE-CR** | Bassetti et al. (2020) | Aged ≥18 years with HAP or VAP or HCAP | 150 (ITT) | Cefiderocol (101) | Best available therapy (61% Colistin-based regimens) (49) | 63.1 ± 19.0/ 63.0 ± 16.7 | 35 vs 66/ 14 vs 35 | 7-14 | 50/26 | 57/21 | 38/ 22 | 15.3 ± 6.5/ 15.4 ± 6.2 | *A. baumannii, K. pneumoniae, P. aeruginosa,* |
| **ATTACK** | Kaye et al. (2023) | Aged ≥18 years with HABP or VABP or VP | 128 (Carbapenem-resistant ABC mMITT) | Sulbactam-durlobactam (64) | Colistin (64) | 62 (54-75)/66 (53-80) | 18 vs 46/ 15 vs 49 | 7-14 | 47/50 | 43/45 | 39/38 | 16 ± 5/ 17 ± 5 | *A. calcoaceticus-baumannii complex* |
| **IMI-REL CHINA** | Li et al. (2024) | Aged ≥18 years with HABP or VABP | 270 (MITT) | Imipenem-cilastatin-relebactam (134) | Piperacillin-tazobactam (136) | 56 ± 15.1/ 59.2 ± 14.7 | 36 vs 98/ 36 vs 100 | 7-14 | 64/56 | 53/54 | NA | 73/75* | *K. pneumoniae, A. calcoaceticus-baumannii complex, P. aeruginosa* |

mean ± SD or median (range), *number of patients in each group with APACHE II score at baseline ≥ 15

ABC = *Acinetobacter baumannii-calcoaceticus*, APACHE II = Acute physiology and chronic health evaluation II, cMITT = clinically modified ITT, F = Female, G = Generic, HABP = Hospital-acquired bacterial pneumonia, HAP = Hospital-acquired pneumonia, HCAP = Healthcare-associated pneumonia, ITT = Intent-to-treat, LRT = Low respiratory tract, M = Male, MITT = modified ITT, mMITT = microbiological modified ITT, N = Newer, VABP = Ventilator-associated bacterial pneumonia, VAP = Ventilator-associated pneumonia, VP = Ventilated pneumonia.

**Supplementary Table 7. Summary of evidence findings**

**Newer antibiotics compared to Generic antibiotics for Hospital-acquired bacterial pneumonia and Ventilator-associated bacterial pneumonia**

**Patient or population:** Hospital-acquired bacterial pneumonia and Ventilator-associated bacterial pneumonia
**Setting:** Intervention: Newer antibiotics; Comparison: Generic antibiotics
**Intervention:** Newer antibiotics; **Comparison:** Generic antibiotics

| Outcomes | № of participants (studies) Follow-up | Certainty of the evidence (GRADE) | Relative effect (95% CI) | Anticipated absolute effects | |
|---|---|---|---|---|---|
| | | | | Risk with Generic antibiotics | Risk difference with Newer antibiotics |
| Day 28 all-cause mortality | 2881 (8 RCTs) | ⊕⊕⊕⊕ High | **RR 0.97** (0.72 to 1.30) | 173 per 1,000 | **5 fewer per 1,000** (48 fewer to 52 more) |
| Favorable clinical response at test-of-cure | 2760 (8 RCTs) | ⊕⊕⊕⊕ High | **RR 1.04** (0.93 to 1.17) | 593 per 1,000 | **24 more per 1,000** (41 fewer to 101 more) |
| Favorable microbiological response at test-of-cure | 1887 (8 RCTs) | ⊕⊕⊕⊕ High | **RR 1.05** (0.89 to 1.24) | 598 per 1,000 | **30 more per 1,000** (66 fewer to 143 more) |
| Nephrotoxicity | 371 (3 RCTs) | ⊕⊕⊕⊕ High | **RR 0.30** (0.11 to 0.79) | 287 per 1,000 | **201 fewer per 1,000** (255 fewer to 60 fewer) |

***The risk in the intervention group** (and its 95% confidence interval) is based on the assumed risk in the comparison group and the **relative effect** of the intervention (and its 95% CI).
**CI:** confidence interval; **RR:** risk ratio

**GRADE Working Group grades of evidence**
**High certainty:** we are very confident that the true effect lies close to that of the estimate of the effect.
**Moderate certainty:** we are moderately confident in the effect estimate: the true effect is likely to be close to the estimate of the effect, but there is a possibility that it is substantially different.
**Low certainty:** our confidence in the effect estimate is limited: the true effect may be substantially different from the estimate of the effect.
**Very low certainty:** we have very little confidence in the effect estimate: the true effect is likely to be substantially different from the estimate of effect.

*Note: GRADE assessment for included RCTs in the systematic review, not only for meta-analysis.

**Supplementary Table 8**. Sensitivity analysis with classic random-effects model in efficacy and safety outcomes

| Meta-analysis Model | Pooled effect estimates | D28ACM | CR | MR | Nephrotoxicity |
|---|---|---|---|---|---|
| Classic random-effects | Pooled RR [95% CI] | 0.96 [0.77; 1.19] | 1.03 [0.95; 1.13] | 1.05 [0.93; 1.18] | 0.30 [0.18; 0.50] |
| | $I_2$ | 30% | 37% | 52% | 0% |

CI = Confidence interval, CR = Clinical response, D28ACM = Day 28 all-cause mortality, $I^2$ = Heterogeneity, MR = Microbiological response, RR = Risk ratio.
